# Data-Efficient Prostate Micro-Ultrasound Image Analysis using a Modality-specific Foundation Model^⋆^

**DOI:** 10.64898/2026.09.29.26364330

**Authors:** Kaifeng Pang, Junxin Fan, Alex Ling Yu Hung, Qi Miao, Changsuk Oh, Qiudi He, Brian Wodlinger, Adam Kinnaird, Wayne Brisbane, Kyunghyun Sung

## Abstract

**Background and Objective:** Micro-ultrasound (MicroUS) is an emerging high-resolution imaging modality for prostate cancer diagnosis and real-time biopsy guidance, but its automated analysis is limited by labor-intensive expert annotation. We aimed to develop and evaluate a foundation model for data-efficient MicroUS image analysis.

**Methods:** We developed *µ*FM, a MicroUS-specific foundation model pretrained using DINOv2-style self-supervised learning on 1,736,391 unlabeled images from 780 patients. The model was fine-tuned and evaluated in three downstream cohorts for prostate capsule segmentation (public, 75 patients), urethra segmentation (institutional, 105 patients), and prostate cancer lesion localization (institutional, 190 patients). Under limited annotation, *µ*FM was compared with randomly initialized models and foundation models pretrained on natural, medical, and ultrasound images. Evaluation used the Dice scores and 95th-percentile Hausdorff distance (HD95) for segmentation and free-response receiver operating characteristic analysis for lesion localization.

**Key Findings and Limitations:** *µ*FM improved performance across all three tasks, with the largest gains under limited annotation. With 10% of training scans, *µ*FM achieved Dice scores of 93.8% for prostate capsule segmentation and 58.4% for urethra segmentation, versus 87.9% and 47.0% for the strongest comparators. Corresponding HD95 values were 2.0 and 4.3 mm. For lesion localization, *µ*FM outperformed other models with approximately 0.75 sensitivity at two false-positive detections per 3D volume. Limitations include retrospective single-institution pretraining and lack of explicit 3D modeling.

**Conclusions and Clinical Implications:** MicroUS-specific self-supervised pretraining improved data-efficient image analysis across prostate and urethra segmentation, and lesion localization. This approach may reduce annotation requirements for developing automated tools in MicroUS image analysis.

**Advancing Practice:** *What does the study add?:* This study evaluates a deep learning model pretrained on a large-scale dataset of unlabeled prostate MicroUS images. The pretrained model was adapted to three clinically relevant image-analysis tasks: prostate capsule segmentation, urethra segmentation, and prostate cancer lesion localization. Substantial performance gains over other models were observed despite limited training data, suggesting that MicroUS-specific self-supervised pretraining may reduce expert annotation requirements for developing automated image-analysis tools.

*Patient Summary:* We developed a foundation model that learns from unlabeled prostate MicroUS images. It performed better with limited labeled data but requires independent validation before clinical use.

## 1. Introduction

Prostate cancer imaging is a central element of diagnosis and risk stratification. For decades, routine imaging and biopsy workflows have been limited to conventional transrectal ultrasound (TRUS) and multiparametric MRI (mpMRI) [1]. The relatively low spatial resolution of conventional TRUS limits direct visualization of suspicious lesions; thus, mpMRI is used to identify target lesions to enable biopsy guidance via modality fusion [2]. While this process is clinically well accepted, it remains susceptible to registration errors between mpMRI and ultrasound [1, 3].

Micro-ultrasound (MicroUS) is an emerging cost-effective 29-MHz ultrasound providing three-to four-fold higher spatial resolution than conventional TRUS [3, 4]. Recent clinical studies indicate that MicroUS-guided biopsy can visualize and detect clinically significant tumors at a rate noninferior to MRI/ultrasound fusion-guided biopsy [3, 5, 6]. The PRI-MUS scores provide a standardized visual risk scale for MicroUS-based prostate cancer diagnosis [7]. However, a consistent critique of MicroUS is that the technology is operator-dependent with acquisition variability and moderate inter-reader variation [8].

Although the OPTIMUM trial provided level 1 evidence supporting MicroUS-guided biopsy, broader clinical adoption will require tools that improve interpretation consistency and reduce annotation burden [5]. AI-based MicroUS analysis has therefore been explored to assist with MicroUS interpretation within tasks such as cancer localization [9, 10], and anatomical segmentation [11, 12]. Existing approaches rely on urologists and radiologists to annotate individual MicroUS images with examples of each task the model is seeking to learn [10, 11]. Some models additionally require anatomical annotations, such as prostate capsule and biopsy needle regions as model inputs [9]. Considering that a single MicroUS scan may contain hundreds of high-resolution slices, such annotation requires multiple hours per case [11], thus limiting scalable model development. In summary, reliable MicroUS models are needed for widespread clinical adoption. MicroUS model development needs a reusable pretrained model that can support multiple MicroUS imaging tasks with limited expert annotation.

Self-supervised foundation models can learn transferable structural and semantic image representations from large-scale unlabeled data [13, 14]. These models can then be fine-tuned with far fewer annotated images for specific downstream applications [13, 14]. However, the effectiveness of downstream fine-tuning depends in part on the similarity between the pretraining and target domains [15]. For example, large-scale visual foundation models such as DINOv2 [14] and iBOT [13] are pretrained mainly on natural images (cats, dogs, etc), while medical foundation models extend pretraining to broad biomedical domains (CT, MRI, US) [16]. More recently, ultrasound-specific foundation models have been developed through large-scale pretraining directly on ultrasound data [17, 18, 19, 20]. Among them, recent work in fetal ultrasound further showed that pretraining on a more narrowly defined domain can outperform representations learned from much larger natural-image datasets, supporting the value of domain-specific foundation models [15]. These findings suggest that progressively more domain-specific pretraining may better capture imaging characteristics relevant to downstream applications. MicroUS represents a specialized imaging domain, with 29-MHz acquisition, fine-grained anatomical texture, and acoustic tissue features associated with prostate cancer appearance [21, 22, 23]. These distinct characteristics may limit the transferability of representations learned from existing pretrained models, motivating MicroUS-specific self-supervised pretraining. In this paper, we developed *µ*FM, a MicroUS-specific foundation model pretrained using DINOv2-style self-supervised learning on approximately 1.7 million unlabeled images from 780 patients. We evaluated the data efficiency of *µ*FM across three downstream tasks: prostate capsule segmentation, urethra segmentation, and prostate cancer lesion localization. For the segmentation tasks, data efficiency was assessed by fine-tuning with 10%, 20%, 50%, and 100% of the available training scans, while lesion localization was evaluated using sparsely annotated lesion-positive slices. We hypothesized that modality-specific pretraining would improve downstream performance when only limited expert annotations were available. To test this hypothesis, we compared *µ*FM with models pretrained on natural-image, medical-image, and ultrasound-image datasets.

## 2. Patients and Methods

### 2.1. Study Design and Data Cohorts

This retrospective study included one large-scale cohort for self-supervised pretraining and three cohorts for downstream evaluation (Table 1). The pretraining, urethra-segmentation, and lesion-localization cohorts were collected at the University of California, Los Angeles (UCLA) Health, while the prostate capsule segmentation cohort comprised a publicly available MicroUS dataset acquired at the University of Florida [11]. For self-supervised pretraining, both MicroUS scans prior to biopsy (initial cine scan) and scans post biopsy were utilized to maximize the scale of unlabeled image data. For downstream evaluation, one pre-biopsy MicroUS scan per patient was included in each cohort. Patients in the downstream evaluation cohorts were excluded from the pretraining cohort at the patient level, ensuring that no imaging data from the same patient were used for both pretraining and downstream evaluation. Training, validation, and test sets were all split at the patient level. The use of UCLA Health data and all experimental studies was approved by the institutional review board (IRB) with a waiver of the requirement for informed consent.

**Table 1:** Overview of the datasets used for self-supervised pretraining and downstream applications.

| Characteristic | Pretraining | Prostate capsule segmentation | Urethra segmentation | Lesion localization |
| --- | --- | --- | --- | --- |
| Data source | Institutional | Public | Institutional | Institutional |
| Patients, $n$ | 780 | 75 | 105 | 190 |
| Scans, $n$ | 6,610 | 75 | 105 | 190 |
| Images, $n$ | 1,736,391 | 2,910 | 2,741 | 6,287 |
| Acquisition period | Jan 2023–Apr 2026 | Sep 2021–Dec 2022 | Sep 2022–Jan 2025 | Jan 2023–Feb 2025 |
| Train/validation/test, patients | – | 45 / 10 / 20 | 60 / 15 / 30 | 110 / 22 / 58* |
| Annotations | None | Prostate capsule masks | Urethra masks | Cancer lesion masks |

\*Lesion-localization splits comprised 95/15, 16/6, and 48/10 positive/negative patients for training, validation, and testing.

### 2.2. MicroUS Image Acquisition and Preprocessing

All MicroUS scans were acquired using the ExactVu MicroUS system (Exact Imaging, Markham, Ontario, Canada). Pre-biopsy anatomy scans were acquired in pseudo-sagittal planes using an imaging depth adjusted to prostate size. Images were converted to grayscale, cropped to remove non-imaging borders, and standardized using statistics estimated from the pre-training cohort. The same preprocessing was applied to all pretraining and downstream datasets. All institutional MicroUS scans and MicroUS-guided biopsies were performed by a single urologist (W.B. - co-author). Additional acquisition and preprocessing details are provided in Supplementary Material S1.

### 2.3. Self-supervised Foundation Model

As shown in Figure 1, *µ*FM was pretrained using a DINOv2-style teacher– student self-distillation framework [14] with DINO [24] and iBOT [13] self-supervised objectives. Both teacher and student encoders used a single-channel ViT-B/16 architecture [25]. Additional model and optimization details are provided in Supplementary Material S2.

**Figure 1:**
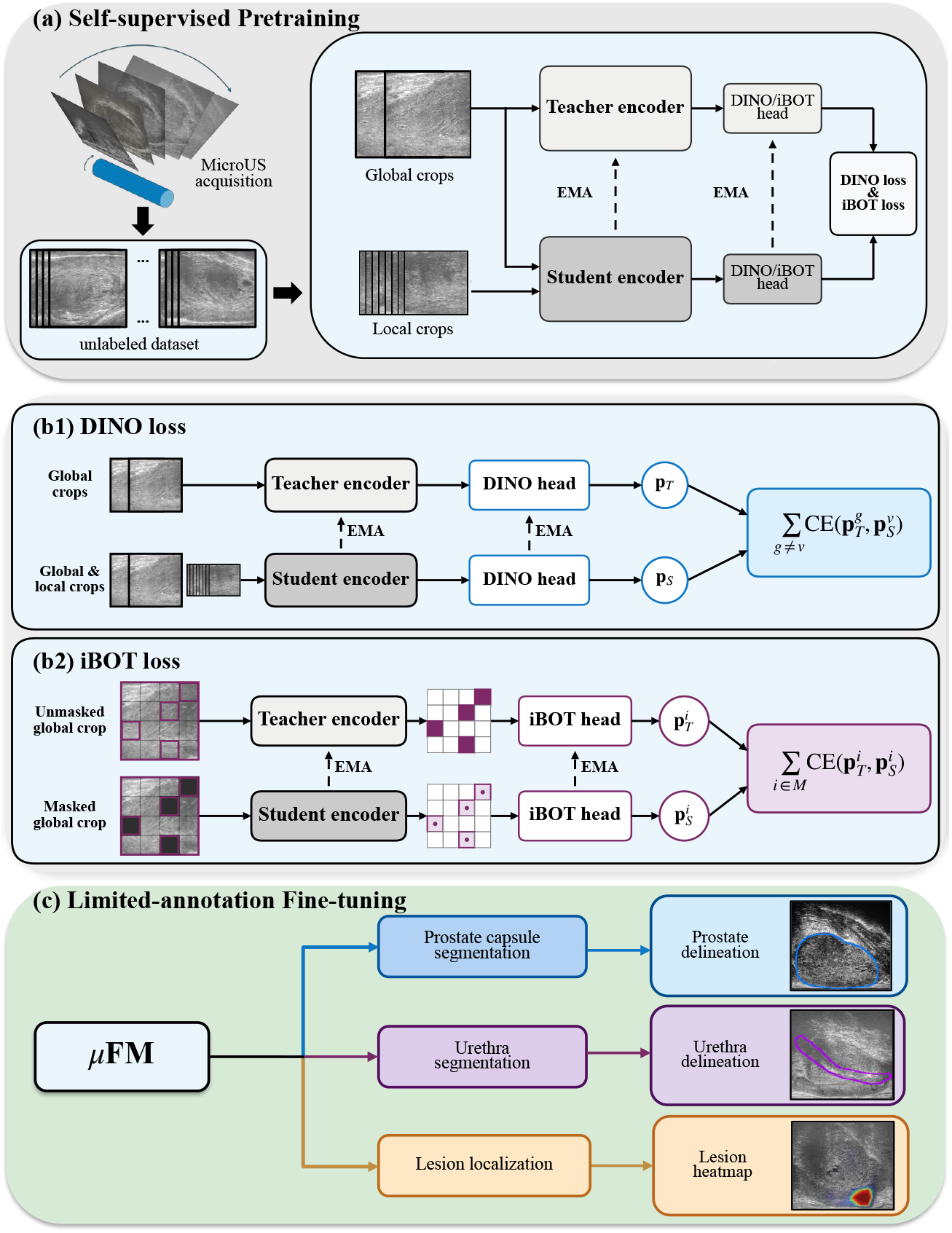
Workflow of *µ*FM. (a) DINOv2 teacher–student self-supervised pretraining on unlabeled MicroUS images. (b1) DINO loss: image-level cross-view objective. (b2) iBOT loss: patch-level objective comparing unmasked teacher targets with predictions at corresponding masked student patches. (c) Limited-annotation fine-tuning for downstream applications.

### 2.4. Downstream Applications and Annotations

We selected three downstream applications to assess clinical performance. Prostate capsule segmentation supports prostate volume assessment and biopsy guidance [11]. Urethra segmentation provides an intraprostatic anatomical landmark relevant to spatial registration and treatment planning [3]. Lesion localization identifies suspicious regions that may support targeted biopsy and diagnosis [10].

For prostate capsule segmentation, we used the public dataset from the University of Florida [11]. For urethra segmentation, one MicroUS scan with the best overall image quality was selected per patient. Reference masks were delineated in OsiriX by an abdominal radiologist (Q.M.) with 2 years of prostate MicroUS interpretation experience and subsequently reviewed and modified when necessary by an experienced MicroUS urologist (W.B.; 6 years of MicroUS experience and *>* 1,000 examinations). For lesion localization, cancer-positive regions were defined by MicroUS-guided targeted biopsy demonstrating clinically significant prostate cancer (csPCa), defined as Grade Group (GG) *≥* 2. Lesion masks were initially delineated by Q.M., guided by the biopsy target location and corresponding visible MicroUS appearance, and subsequently reviewed by W.B. Training cases were sparsely annotated using 1–6 lesion-positive images per positive patient, whereas all lesion-positive images were annotated for validation and testing. Biopsy-negative patients were also included. Cohort sizes and patient-level splits are summarized in Table 1. Additional data and annotation details are provided in Supplementary Material S3.

### 2.5. Limited-Annotation Fine-Tuning and Comparison Models

To evaluate data efficiency, prostate capsule and urethra segmentation models were fine-tuned using 10%, 20%, 50%, and 100% of the available training scans, with data sub-sampled at the scan level. For lesion localization, the sparsely annotated training cohort was used without additional scan-level subsampling. We compared *µ*FM with encoders pretrained on natural images (iBOT [13] and DINOv2 [14]), biomedical images (BiomedCLIP [16]), and ultrasound images (USFM [17], URFM [18], USF-MAE [20], and EchoCare [19]). Non-pretrained comparators included nnUNet [26], TransUNet [27], MicroSegNet [11], and a randomly initialized ViT-B [25]. All experiments were repeated five times with different random seeds. Additional implementation details are provided in Supplementary Material S4.

### 2.6. Evaluation Metrics and Statistical Analysis

For prostate capsule and urethra segmentation, slice-wise predictions were aggregated into 3D volumes and evaluated using the Dice similarity coefficient (DSC) and 95th-percentile Hausdorff distance (HD95). For each patient, metrics were averaged across five runs, and *µ*FM was compared with other methods using two-sided Wilcoxon signed-rank tests with Holm correction. Lesion localization was evaluated using free-response receiver operating characteristic (FROC) curves of lesion-level sensitivity versus false positives per volume, averaged across five runs. Additional evaluation details are provided in Supplementary Material S5.

## 3. Results

### 3.1. Prostate Capsule Segmentation

As shown in Table 2.A, *µ*FM achieved the highest DSC and lowest HD95 across all annotation settings in prostate capsule segmentation. With only 10% of the training scans used for fine-tuning, *µ*FM achieved a DSC of 93.8 *±* 0.6% and an HD95 of 2.0 *±* 0.4 mm, improving DSC by 5.9 percentage points and reducing HD95 by 3.8 mm compared with the strongest pretrained competitor, EchoCare. Paired two-sided Wilcoxon signed-rank tests with Holm correction showed that *µ*FM significantly outperformed the comparison methods at the 10%, 20%, and 50% annotation settings (*p*_Holm_ *<* 0.05).

**Table 2:** Quantitative comparison of prostate capsule and urethra segmentation under limited-annotation settings: (A) Prostate capsule segmentation, and (B) Urethra segmentation. Results are reported as mean *±* standard deviation across five runs and were computed at the 3D volume level by aggregating slice-wise predictions. DSC = Dice similarity coefficient; HD95 = 95th-percentile Hausdorff distance.

| Training Ratio | 10% |  | 20% |  | 50% |  | 100% |  |
| --- | --- | --- | --- | --- | --- | --- | --- | --- |
| Baselines | DSC (%) ↑ | HD95 (mm) ↓ | DSC (%) ↑ | HD95 (mm) ↓ | DSC (%) ↑ | HD95 (mm) ↓ | DSC (%) ↑ | HD95 (mm) ↓ |
| **A. Prostate capsule segmentation** |  |  |  |  |  |  |  |  |
| nnUNet | 85.8±1.3 | 7.5±0.5 | 89.1±1.1 | 5.6±0.6 | 93.3±0.6 | 2.7±0.5 | 94.6±0.1 | 1.9±0.1 |
| TransUNet | 72.0±2.2 | 10.0±0.4 | 75.6±2.1 | 8.9±0.9 | 82.1±1.4 | 8.0±0.4 | 82.8±0.3 | 7.9±0.2 |
| MicroSegNet | 85.8±1.9 | 7.4±1.1 | 89.5±1.0 | 5.6±0.8 | 91.8±0.5 | 4.5±0.5 | 93.4±0.3 | 3.5±0.4 |
| ViT | 80.6±1.9 | 8.3±0.9 | 83.3±0.5 | 7.8±0.4 | 88.4±0.5 | 6.2±0.5 | 90.8±0.3 | 5.1±0.6 |
| ViT+iBOT | 80.3±1.4 | 7.7±0.5 | 82.7±0.8 | 7.5±0.2 | 86.9±0.8 | 6.4±0.3 | 89.8±0.2 | 5.5±0.1 |
| ViT+DINOv2 | 81.3±0.6 | 7.6±0.5 | 83.5±0.7 | 7.3±0.3 | 87.9±0.8 | 5.9±0.5 | 90.8±0.7 | 5.0±0.7 |
| ViT+BiomedCLIP | 79.1±1.9 | 8.3±1.0 | 82.8±0.9 | 7.2±0.2 | 87.4±0.3 | 6.2±0.2 | 89.7±0.2 | 5.4±0.2 |
| ViT+USFM | 80.2±2.4 | 8.2±0.9 | 84.0±0.9 | 6.7±0.6 | 88.6±0.7 | 5.7±0.4 | 91.3±0.2 | 4.1±0.1 |
| ViT+URFM | 82.6±1.7 | 7.3±0.7 | 85.9±0.2 | 6.4±0.6 | 89.1±0.9 | 5.8±0.6 | 91.8±0.3 | 4.1±0.6 |
| ViT+USF-MAE | 81.1±1.5 | 8.1±0.8 | 84.4±0.8 | 7.2±0.7 | 88.8±0.2 | 6.1±0.5 | 91.9±0.5 | 3.9±0.5 |
| ViT+EchoCare | 87.9±1.3 | 5.8±1.3 | 90.2±1.4 | 4.3±0.9 | 93.1±0.6 | 2.9±0.7 | 93.9±0.4 | 2.4±0.1 |
| **ViT+μFM** | **93.8±0.6** | **2.0±0.4** | **94.3±0.2** | **1.8±0.2** | **94.8±0.2** | **1.6±0.1** | **95.1±0.1** | **1.5±0.04** |
| **B. Urethra segmentation** |  |  |  |  |  |  |  |  |
| nnUNet | 41.9±3.9 | 7.9±0.5 | 41.3±3.9 | 7.5±0.3 | 51.6±1.1 | 5.6±0.2 | 54.4±2.9 | 4.9±0.3 |
| TransUNet | 31.1±2.8 | 10.2±0.7 | 33.5±1.3 | 10.4±0.2 | 38.1±2.3 | 9.2±0.4 | 40.4±2.5 | 8.4±0.5 |
| MicroSegNet | 42.6±1.8 | 9.1±0.9 | 44.9±4.4 | 7.5±1.3 | 53.0±0.5 | 7.2±0.5 | 58.2±1.1 | 5.9±0.6 |
| ViT | 41.4±3.0 | 7.6±1.1 | 42.4±4.2 | 6.8±1.0 | 50.2±1.1 | 5.4±0.2 | 52.8±1.7 | 5.1±0.4 |
| ViT+iBOT | 42.3±3.5 | 7.6±0.7 | 43.8±3.5 | 7.2±0.8 | 50.2±1.5 | 6.0±0.9 | 54.6±0.9 | 4.5±0.2 |
| ViT+DINOv2 | 39.9±2.4 | 8.3±0.8 | 41.3±4.3 | 7.4±0.6 | 48.1±3.2 | 5.4±0.4 | 50.9±1.6 | 4.9±0.3 |
| ViT+BiomedCLIP | 42.2±2.6 | 7.2±0.6 | 43.4±1.6 | 6.8±0.5 | 50.9±1.4 | 5.4±0.7 | 54.7±1.9 | 4.5±0.1 |
| ViT+USFM | 40.4±1.5 | 7.7±0.4 | 42.5±3.4 | 7.3±0.5 | 46.8±2.9 | 6.1±0.4 | 49.3±0.9 | 5.6±0.6 |
| ViT+URFM | 43.3±2.0 | 6.8±0.1 | 45.3±2.7 | 6.9±1.0 | 53.6±2.5 | 4.9±0.6 | 57.5±1.4 | 4.4±0.2 |
| ViT+USF-MAE | 42.5±2.8 | 7.7±0.8 | 43.7±3.4 | 6.8±0.4 | 50.1±2.5 | 5.2±0.2 | 56.2±1.2 | 4.6±0.2 |
| ViT+EchoCare | 47.0±3.7 | 6.8±0.3 | 47.1±3.6 | 6.7±0.4 | 55.3±0.9 | 5.4±0.5 | 59.4±1.4 | 4.8±0.3 |
| **ViT+μFM** | **58.4±1.0** | **4.3±0.2** | **60.0±1.6** | **4.4±0.5** | **62.8±0.7** | **3.7±0.4** | **63.1±0.2** | **3.4±0.1** |

At the 100% data setting, differences from nnUNet were not statistically significant for DSC (*p*_Holm_ = 0.648) or HD95 (*p*_Holm_ = 0.059). Representative qualitative comparisons are shown under the 10% annotation setting in Figure 2.A, including MicroSegNet as the best-performing non-pretrained model and EchoCare as the best-performing pretrained baseline. Compared with these methods, *µ*FM produced more accurate gland boundaries, fewer false-positive segmentations, and improved segmentation continuity.

**Figure 2:**
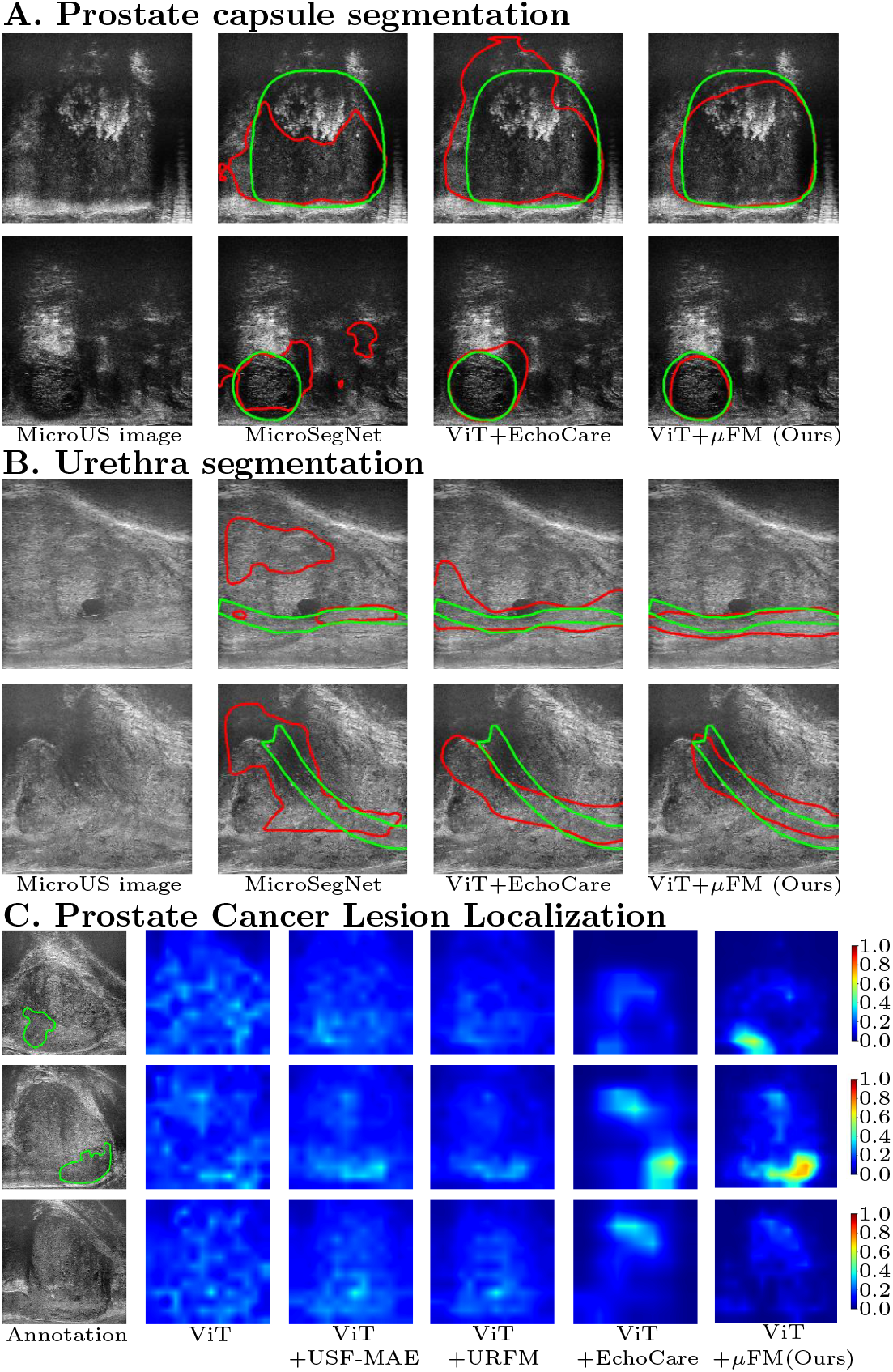
Representative qualitative results for downstream applications. Predictions from the best-performing run of each method are shown. (A) Prostate capsule and (B) urethra segmentation under the 10% annotation setting, with reference contours in green and predictions in red. (C) Prostate cancer lesion localization in two biopsyconfirmed csPCa cases and one biopsy-negative case. Reference lesion contours are shown in green for positive cases, and warmer colors in the probability maps indicate higher predicted probabilities of csPCa.

### 3.2. Urethra Segmentation

For urethra segmentation, *µ*FM achieved the highest DSC and lowest HD95 across all annotation settings (Table 2.B). With only 10% of the training scans used for fine-tuning, *µ*FM achieved a DSC of 58.4 *±* 1.0% and an HD95 of 4.3 *±* 0.2 mm, improving DSC by 11.4 percentage points over EchoCare and reducing HD95 by 2.5 mm compared with URFM. In paired comparisons using two-sided Wilcoxon signed-rank tests with Holm correction, *µ*FM significantly outperformed all comparison methods for HD95 across all annotation settings. For DSC, *µ*FM significantly outperformed all comparison methods at the 10%, 20%, and 50% annotation settings. With 100% of the training data, *µ*FM remained significantly better than all methods except MicroSegNet (*p*_Holm_ = 0.839) and EchoCare (*p*_Holm_ = 0.063). Representative 10% annotation results are shown in Figure 2.B, where *µ*FM produced more continuous and anatomically coherent urethral delineations than the selected comparators.

### 3.3. Prostate Cancer Lesion Localization

As shown in Figure 3, *µ*FM achieved higher lesion-level sensitivity than the comparison methods across most of the evaluated false-positive range. At 1 false-positive detection per volume, *µ*FM achieved a sensitivity of 0.648 *±* 0.023, compared with 0.507 *±* 0.026 for URFM, the strongest competitor. At 2 false-positive detections per volume, *µ*FM achieved a sensitivity of 0.762 *±* 0.030, compared with 0.559 *±* 0.018 for URFM.

**Figure 3:**
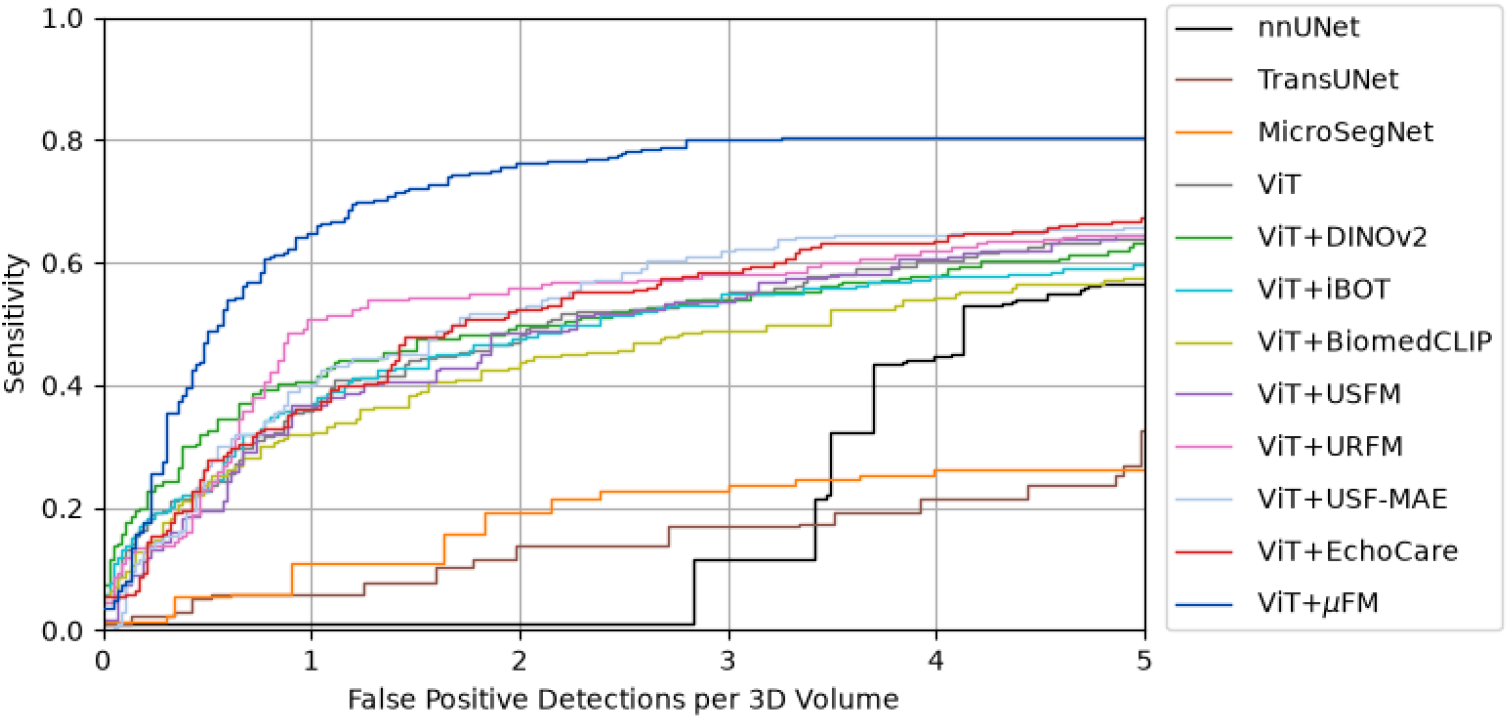
Free-response receiver operating characteristic (FROC) curves for prostate cancer lesion localization. Curves represent the point-wise average across five runs.

Representative qualitative results are shown in Figure 2.C, including the randomly initialized ViT and strong pretrained baselines: USF-MAE, URFM, and EchoCare. Qualitatively, in the representative cases shown in Figure 2.C, *µ*FM produced more spatially concentrated probability responses that corresponded more closely to the reference lesions in the two biopsy-confirmed csPCa cases and showed fewer false-positive responses in the biopsy-negative case.

## 4. Discussion

We developed *µ*FM, a MicroUS-specific foundation model using large-scale self-supervised pretraining. *µ*FM consistently improved performance across all three downstream tasks: prostate capsule segmentation, urethra segmentation, and prostate cancer lesion localization. *µ*FM outperformed non-pretrained models and models pretrained on natural-image, medical-image, and ultrasound datasets. For both segmentation tasks, statistically significant improvements were observed across most annotation settings, with particularly large gains when only a small proportion of the available training scans were used for fine-tuning. For lesion localization, *µ*FM achieved higher lesion-level sensitivity across most of the evaluated false-positive range despite sparse training annotations. These findings support MicroUS-specific self-supervised pretraining as a data-efficient strategy for MicroUS image analysis.

For MicroUS imaging, a single clinical scan may contain hundreds of high-resolution images, making dense expert annotation labor-intensive and hindering the development of automated image-analysis tools. Our limited-annotation experiments showed that MicroUS-specific pretraining can reduce the task-specific annotation burden while providing transferable representations across different downstream applications. With only 10% of training scans used for fine-tuning, *µ*FM achieved a prostate capsule segmentation DSC of 93.8%, approaching the 94.6% achieved by nnUNet using the full training cohort, while for urethra segmentation it achieved a DSC of 58.4%, comparable to or higher than all competing methods trained using all training scans. This benefit was particularly evident for urethra segmentation, which is more challenging than prostate capsule segmentation due to anatomic variation, smaller size, and limited visibility across MicroUS slices. Despite this increased difficulty, *µ*FM maintained the best performance across all annotation settings, suggesting that modality-specific pretraining may be particularly valuable for anatomically challenging tasks with limited supervision. The narrowing performance gap as more labeled data became available further suggests that the primary benefit of self-supervised pretraining lies in improving data efficiency. Moreover, the favorable performance of *µ*FM for prostate cancer lesion localization extends its transferability beyond anatomical segmentation to a more complex diagnosis-related task, where models must identify subtle and heterogeneous patterns associated with biopsy-confirmed csPCa while minimizing false-positive detections. Despite annotating only a few lesion-positive slices per patient for training, *µ*FM achieved higher lesion-level sensitivity than competing methods across most of the evaluated free-response receiver operating characteristic (FROC) range, including at low false-positive burdens. Previous MicroUS studies have demonstrated the feasibility of AI-based prostate cancer localization and detection using task-specific models or pretrained representations [10, 28]. Our findings complement these studies by showing that a MicroUS-specific pretrained representation can support lesion localization with sparse annotations while also transferring to anatomical segmentation tasks.

Our results also highlight the value of domain-specific pretraining for MicroUS, as *µ*FM consistently outperformed models pretrained on natural-image, medical-image, and conventional ultrasound datasets. Although existing foundation models provide broadly transferable representations [14, 16, 18], MicroUS has distinct imaging characteristics, including fine-grained anatomical texture, specific acoustic patterns for cancer and benign tissue [21, 22, 23], and pseudo-sagittal acquisition, which may be better captured by pretraining directly on MicroUS images than by transferring representations learned from less related domains. Our experimental findings are consistent with prior work showing benefits from domain-specific self-supervised pretraining in ultrasound imaging [15, 19]. Importantly, the advantage of *µ*FM over broader ultrasound-pretrained models [17, 18, 19, 20] suggests that pretraining within the prostate MicroUS domain may provide additional benefits beyond ultrasound-specific pretraining alone.

Despite the promising performance of *µ*FM, several limitations remain. First, the pretraining and in-house downstream cohorts were retrospectively collected at a single institution using the same MicroUS platform and were acquired by the same operator. Although prostate capsule segmentation was evaluated on an independently acquired public dataset, urethra segmentation and lesion localization have not yet been validated prospectively or in independent external cohorts, which may limit generalizability across institutions, operators, and acquisition settings. Second, although performance was evaluated at the 3D volume level by aggregating slice-wise predictions, *µ*FM operates on 2D images and does not explicitly model inter-slice spatial information. Third, the study evaluated algorithmic performance rather than clinical deployment, and the impact of *µ*FM on biopsy targeting, workflow efficiency, or clinical decision-making requires prospective evaluation. Future studies will evaluate *µ*FM across multiple institutions, operators, and acquisition settings. We will also explore 3D volumetric modeling and prospectively evaluate its utility within clinical MicroUS workflows.

## 5. Conclusions

We developed *µ*FM, a MicroUS-specific foundation model pretrained using large-scale self-supervised learning on unlabeled prostate MicroUS images. *µ*FM consistently improved performance across prostate capsule segmentation, urethra segmentation, and prostate cancer lesion localization, with the largest improvements observed under limited-annotation settings. These findings demonstrate that MicroUS-specific pretraining provides a data-efficient and transferable strategy by reducing task-specific annotation requirements while maintaining strong downstream performance across multiple MicroUS image-analysis tasks. Further prospective and multi-institutional evaluation will be conducted to establish its utility in clinical MicroUS work-flows.

## Data Availability

The data used in this study are not publicly available
due to patient privacy and institutional restrictions, but may
be available from the corresponding author upon reasonable
request and with permission from the Institutional Review
Board (IRB).

## Take Home Message

MicroUS-specific self-supervised pretraining reduced annotation requirements while improving prostate capsule segmentation, urethra segmentation, and prostate cancer lesion localization.

