## Supplementary Material for "Data-Efficient Prostate Micro-Ultrasound Image Analysis using a Modality-specific Foundation Model^⋆^"

### S1. MicroUS Image Acquisition and Preprocessing

All MicroUS scans used in our study were acquired using the ExactVu MicroUS system (Exact Imaging, Markham, Ontario, Canada). An initial "anatomy scan" was acquired before the biopsy to minimize biopsy-related imaging artifacts. The prostate was manually scanned in pseudo-sagittal planes, with an imaging depth of 3–6 cm adjusted according to prostate size to ensure coverage of the entire gland. The probe was bound by the rectum under the prostate, and scans were obtained by rotating the probe from right to left, with the piezoelectric crystals contacting the anterior rectum directly adjacent to the posterior prostate [1]. All institutional MicroUS scans and MicroUS-guided biopsies were performed by a single urologist (W.B. - co-author).

Each prostate cine scan contained up to 300 MicroUS images, with the angular field of view varying from 109.3° to 185.4° depending on patient anatomy. Larger prostates required a larger field of view. The acquired images had a matrix size of $962 \times 1372$ pixels and an in-plane pixel spacing of 70 $\mu$m. As a preprocessing step before model development, all images were

*Corresponding author.
[1] These authors contributed equally to this work.

converted to grayscale and cropped to remove non-imaging black borders. Pixel intensities were standardized using the mean and standard deviation estimated from the pretraining cohort. The same preprocessing was applied to the pretraining and downstream datasets before model training and evaluation.

### S2. Self-supervised Foundation Model

$\mu$FM was pretrained using a DINOv2-style teacher–student self-distillation framework [2] with two self-supervised objectives, DINO [3] and iBOT [4]. Both teacher and student encoders used the same single-channel ViT-B/16 architecture [5]. For each image, two global crops ($224 \times 224$) and eight local crops ($96 \times 96$) were randomly extracted. For the DINO objective, the teacher generated image-level predictions from the two global crops, while the student generated predictions from both global and local crops. The DINO objective encouraged predictions from different views of the same image to be consistent between the teacher and student networks. The iBOT objective operated only on the global crops. Sampled patch positions in the student global crops were replaced with learnable mask tokens, while the corresponding unmasked teacher patches served as targets for masked patch prediction. Both DINO and iBOT losses used the cross-entropy (CE) loss function. The student network was optimized by backpropagation, while the teacher parameters were updated using an exponential moving average (EMA) of the student parameters. The global and local crop area fractions were sampled from 0.5–1.0 and 0.2–0.32, respectively. Pretraining was performed for 50,000 iterations using the AdamW optimizer [6], with a global batch size of 288 across three NVIDIA H100 GPUs.

### S3. Downstream Applications and Annotations

#### *S3.1. Prostate capsule segmentation*

For prostate capsule segmentation, we used the public dataset from the University of Florida [7]. Prostate capsule masks were initially annotated by two non-expert annotators and iteratively reviewed by a urologist with MicroUS expertise until consensus was reached. These expert-reviewed masks were used as reference annotations. The original dataset included 55 training and 20 test patients. We further split the training cohort into 45 training and 10 validation patients while retaining the original 20-patient test cohort.

#### *S3.2. Urethra segmentation*

For urethra segmentation, one MicroUS scan with the best overall image quality was selected for each patient. Reference masks were delineated in OsiriX by an abdominal radiologist (Q.M.) with 2 years of prostate MicroUS interpretation experience and subsequently reviewed and modified when necessary by an experienced MicroUS urologist (W.B.; 6 years of MicroUS experience and $>$ 1,000 examinations). Urethral contours were delineated on images in which the urethra could be reliably identified, with the bladder neck, verumontanum, and bulbar urethra visible in a single sagittal plane. The cohort was randomly divided at the patient level into 60 training, 15 validation, and 30 test patients.

#### *S3.3. Prostate cancer lesion localization*

For lesion localization, cancer-positive regions were defined by MicroUS-guided targeted biopsy demonstrating clinically significant prostate cancer (csPCa), defined as Grade Group (GG) $\geq 2$. Using biopsy histopathology as the reference standard, lesion delineation was guided by the biopsy target location and corresponding visible lesion appearance on MicroUS. Masks were initially delineated by Q.M. and subsequently reviewed and modified when necessary by W.B. The training cohort was sparsely annotated with 1–6 lesion-positive images per positive patient, selected from visually apparent portions near the beginning, middle, and end of the lesion extent. In the validation and test cohorts, all lesion-positive images were annotated, resulting in 7–177 annotated images per positive patient. Thirty-one biopsy-negative patients were additionally included as negative cases and distributed across the training, validation, and test sets as 15, 6, and 10 patients, respectively. For negative cases, 20 images were randomly sampled from each training scan, while the central 100 consecutive images were selected for validation and testing.

### S4. Limited-Annotation Fine-Tuning and Comparison Models

To evaluate data efficiency, we conducted limited-annotation fine-tuning for the three downstream applications. For prostate capsule and urethra segmentation, models were fine-tuned using 10%, 20%, 50%, and 100% of the available training scans, with training data sub-sampled at the scan level. For lesion localization, the sparsely annotated training cohort was used directly without additional scan-level subsampling. We compared $\mu$FM with natural-image pretrained encoders (iBOT [4] and DINOv2 [2]), a biomedical-image pretrained encoder (BiomedCLIP [8]), and ultrasound-specific pretrained encoders (USFM [9], URFM [10], USF-MAE [11], and EchoCare [12]). Non-pretrained comparators were additionally included, i.e., nnUNet [13], TransUNet [14], MicroSegNet [7], and a randomly initialized ViT-B [5]. Official ViT-B-scale checkpoints were used for all pretrained encoders. The randomly initialized ViT-B used the same decoder architecture and training configuration as the pretrained ViT-based models.

During fine-tuning, images were resized to $224 \times 224$ using bilinear interpolation and standardized using pretraining-cohort statistics. For segmentation, a lightweight CNN decoder [15] was attached to each pretrained encoder. Prostate capsule segmentation used annotation-guided binary cross-entropy loss [7], whereas urethra segmentation used equally weighted Dice and binary cross-entropy losses. For lesion localization, a $1 \times 1$ convolutional decoder was used with focal loss [16] ($\gamma = 2.0$, $\alpha = 0.9$). All pretraining-based models were optimized using AdamW [6] with a batch size of 8 and learning rates of $5 \times 10^{-5}$ and $1 \times 10^{-3}$ for the encoder and decoder, respectively. Segmentation and lesion localization models were trained for 100 and 30 epochs, respectively. All fine-tuning experiments were repeated five times with different random seeds for training-set subsampling and model initialization.

### S5. Evaluation Metrics and Statistical Analysis

For prostate capsule and urethra segmentation, slice-wise predictions were aggregated into 3D volumes and evaluated using the Dice similarity coefficient (DSC) and 95th-percentile Hausdorff distance (HD95). Patient-level metrics were averaged across five runs, and paired differences between $\mu$FM and other methods were assessed using two-sided Wilcoxon signed-rank tests with Holm correction. For lesion localization, slice-wise probability maps were aggregated into 3D volumes. Following prior studies [17, 18, 19], candidate detections were identified as local probability maxima within a 5-mm-radius neighborhood and considered true positives if located within a reference lesion or within 0.5 mm of its boundary. Performance was assessed using free-response receiver operating characteristic (FROC) curves of lesion-level sensitivity versus false positives per volume, computed for each run and averaged point-wise across five runs.
